# Generating Evidence for Chronic Obstructive Pulmonary Disease (COPD) Clinical Guidelines Using EHR Data

**DOI:** 10.1101/19006023

**Authors:** Amber M. Johnson, Marvi Bikak, Paul M. Griffin, Mohammad Adibuzzaman

## Abstract

**Objectives:** The aim of this research was to develop data-driven models using electronic health records (EHRs) to conduct clinical studies for predicting clinical outcomes through probabilistic analysis that considers temporal aspects of clinical data. We assess the efficacy of antibiotics treatment and the optimal time of initiation for in-hospitalized diagnosed with acute exacerbation of COPD (AECOPD) as an application to probabilistic modeling.

**Materials and Methods:** We developed a semi-automatic Markov Chain Monte Carlo (MCMC) modeling and simulation approach that encodes clinical conditions as computable definitions of health states and exact time duration as input for parameter estimations using raw EHR data. We applied the MCMC approach to the MIMIC-III clinical database, where ICD-9 diagnosis codes (491.21, 491.22, and 494.1) were used to identify data for 697 AECOPD patients of which 25.9% were administered antibiotics.

**Results:** The average time to antibiotic administration was 27 hours, and 32% of patients were administered *vancomycin* as the initial antibiotic. The model simulations showed a 50% decrease in mortality rate as the number of patients administered antibiotics increased. There was an estimated 5.5% mortality rate when antibiotics were initially administrated after 48 hours vs 1.8% when antibiotics were initially administrated between 24 and 48 hours.

**Discussion:** Our findings suggest that there may be a mortality benefit in initiation of antibiotics early in patient with severe respiratory failure in settings of COPD exacerbations warranting an ICU admission.

**Conclusion:** Probabilistic modeling and simulation methods that considers temporal aspects of raw clinical patient data can be used to adequately generate evidence for clinical guidelines.

## BACKGROUND AND SIGNIFICANCE

Clinical guidelines are systematically developed documents that support patient care for specific clinical circumstances.^1^ They are typically developed through either evaluation of the “best” available evidence generated via meta-analyses, randomized controlled trials, or expert opinion.^2^ Such evidence-based sources supplement physician experience, which is typically based on individual clinical experience and discussion with other physicians.^3,4^ In practice, physicians may not follow the clinical guidelines due to a lack of awareness, conflicting recommendations from the presence of different clinical conditions in a patient, or limited recommendations due to lack of evidence.^5^ For example, the Global Initiative for Chronic Obstructive Lung Disease (GOLD) national clinical practice guidelines^6^ have been used worldwide by healthcare professionals for treating and managing chronic obstructive pulmonary disease (COPD). COPD refers to a group of diseases including chronic bronchitis and emphysema, that effects over 16 million persons in the U.S., and is the third leading cause of death.^7^ GOLD recommends antibiotic therapy for patients with severe acute exacerbation of COPD (AECOPD), a sudden worsening of symptoms,^8^ as they can shorten hospital length-of-stay (LOS) and decrease mortality. Though several studies^9^ have been conducted to assess the short-term and long-term efficacy of antibiotics for AECOPD none have explored the timing of when antibiotics are administered to AECOPD patients in the intensive care unit (ICU). Hence, there are no guidelines or recommendations for the initial timing for administering antibiotics.

AECOPD often requires multiple hospitalizations over a patient’s lifetime. During these hospitalizations physician care decisions are collected in patients’ electronic health records (EHRs) and clinical notes. Each visit is a source of patient information (e.g., diagnosis, treatment, outcome) about clinical events that effect their health state during their hospital encounters (i.e., care pathway). While this information is crucial for treating patients, there is often not a standardized method of processing and documenting the information.^10^ Moreover, data can be challenging to retrieve and properly interpret due to its high volume and unstructured nature.^11^ Existing software systems that analyze EHR data^12,13^ do not allow for longitudinal processes like chronic disease progression to be observed directly, nor do they support analysis of how patients transition from one health state to another as a direct cause of an intervention (e.g., drug administration, oxygen therapy, surgery).

Despite the limitations of current tools, there are examples of retrospective data in EHRs being used for evidence generation through longitudinal analysis.^13–15^ This process helps identify medical trends for a patient population and improve the overall quality of care, which is particularly important for chronic conditions. Markov models are one approach that have been used to estimate healthcare costs, utilization, and disease progression over time.^16–18^ They provide support for evaluating uncertainties in decision making by approximating patient transitions through a set of “health states”, each of which corresponds to a clinical event.^19^ While using Markov chains has been useful for chronic conditions such as COPD, directly using EHR data in Markov models presents certain challenges. In particular, Markov chains assume a one-step time-invariant transition. EHRs, however, contain highly-dimensional, time-variant, clinical data observed at irregular time intervals. Hence, the nature of EHR data limits the ability to represent granular timing of the transitions, which can lead to process misrepresentation in the model. Thus, EHR data points must be transformed and common modeling parameter estimation techniques refined to develop Markov models that properly capture temporal characteristics.

This research presents an approach that systematically captures the effect of interventions during medical encounters, and hence supports the development of clinical guidelines. A semi-automatic Markov Chain Monte Carlo (MCMC) modeling and simulation approach that encodes clinical conditions as computable definitions of health states using raw EHR data is described. We further demonstrate how data-driven models can be used to conduct clinical studies to predict clinical outcomes through probabilistic analysis. While our approach is general, we present an illustration by considering the efficacy of antibiotics treatment for in-hospitalized AECOPD patients. A model of the pathway of AECOPD is generated for hospitalized patients over time by curating retrospective EHR data collected during critical care encounters. Recommendations from the national GOLD guidelines^6^ and physicians were incorporated to define model components. We estimated outcomes for two research questions related to antibiotic treatment for AECOPD patients, namely: i) the impact of antibiotic administration on in-hospital death and ii) the impact on in-hospital death based on the initial timing of antibiotic administration. We performed Monte Carlo simulations, which we validated by comparing the estimated model outcomes to actual outcomes from the EHR data, and conducted sensitivity analyses.

## MATERIALS AND METHODS

### Data

Data was used from MIMIC-III (Medical Information Mart for Intensive Care),^20,21^ a curated database from Beth Israel Deaconess Medical Center, comprised of nearly 40,000 distinct patients and nearly 50,000 hospital admissions for adult patients in critical care units from 2001 to 2012. Each patient visit was associated with multiple diagnoses, which were labeled by the International Classification of Diseases, Ninth Revision (ICD-9) diagnosis codes and ordered by priority. We defined a primary diagnosis of AECOPD as a hospital admission having at least one of the following ICD-9 codes as either primary, secondary or tertiary diagnosis:^22,23^ 491.21, 491.22, and 494.1. We limited our data extraction to these three codes since we wanted to analyze patients with acute respiratory failure (ARF) secondary to AECOPD primarily, without other confounding etiologies that may be present. Based on this selection criteria (Figure 1), we identified 697 unique AECOPD patients with at least one ICU admission.

**Figure 1:**
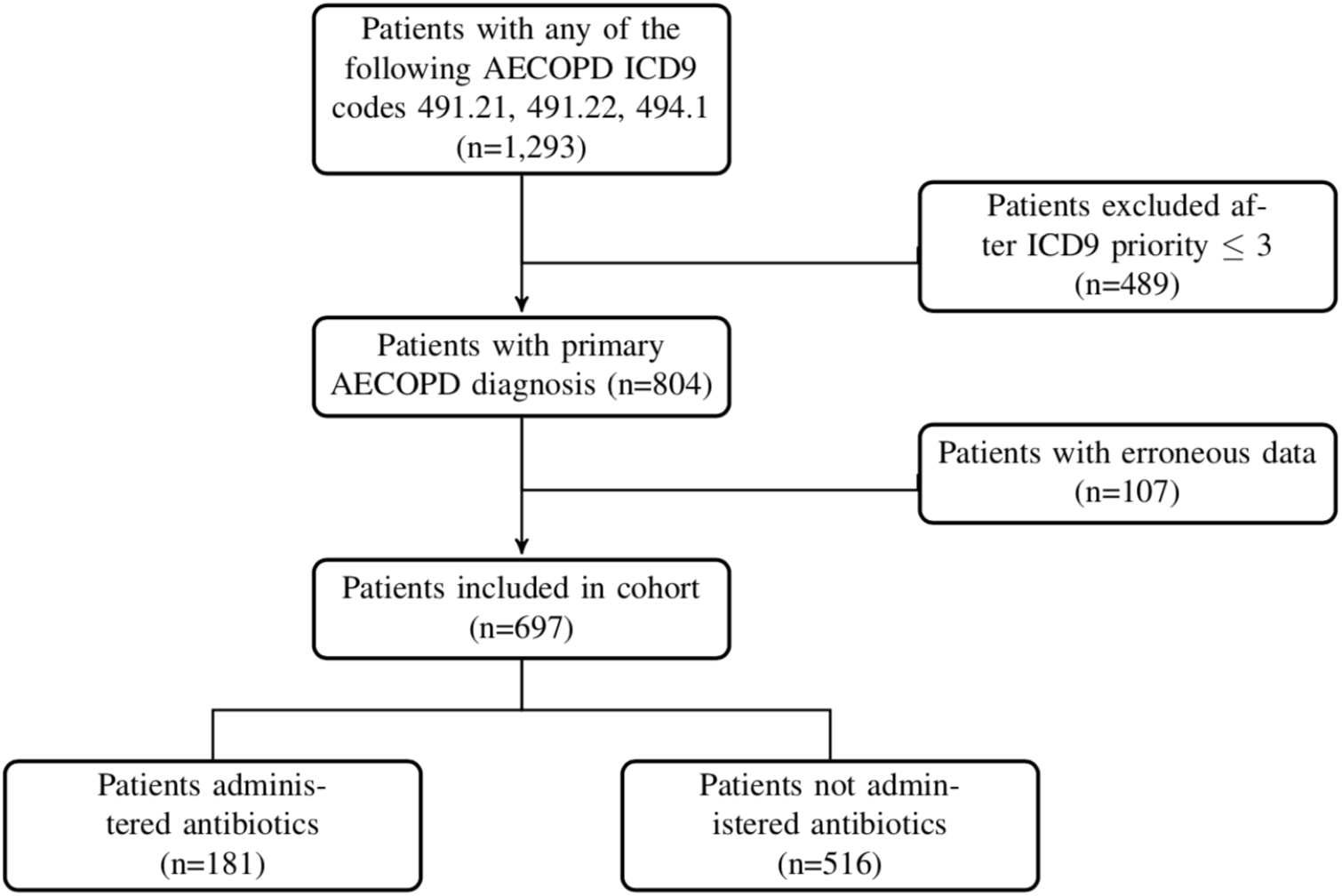
Cohort selection criteria for AECOPD patients administered and not administered antibiotics in the MIMIC-III database.

GOLD guidelines suggest that severe exacerbations requiring hospitalization may be associated with ARF,^6^ a build up in the air sac of the lungs that inhibits the release of oxygen into the blood.^8^ There are two types of ARF, hypoxemic (type 1) and hypercapnic (type 2), which can be identified by changes in clinical variables that GOLD recommends for classifying exacerbations^6^ are: partial pressure of oxygen (PaO_2_), partial pressure of carbon dioxide (PaCO_2_), and pH. We used recommendations from GOLD and consulted with COPD experts to identify and define health states on the basis of established clinical criteria^6^ in terms of these variables (Table 1). We extracted the time that a clinical measurement or event was recorded and determined the health state for the patient at that time to create temporal paths, represented by pairs of ordered time-stamps and state sequences.

**Table 1:** Description of health and outcome states.

| State Identifier |  | Definition | State Type |
| --- | --- | --- | --- |
| 0 | NARF | No acute respiratory failure | Health |
| 1 | ARF<sub>1</sub> | PaO<sub>2</sub> ≤ 60<br>PaCO<sub>2</sub> ≤ 50 | Health |
| 2 | ARF<sub>2</sub> | PaCO<sub>2</sub> > 50<br>pH ≤ 7.25<br>PaO<sub>2</sub> ≤ 60<br>PaCO<sub>2</sub> > 50<br>pH ≤ 7.25 | Health |
| 3 | Discharge | Discharge from hospital | Outcome |
| 4 | Death | In-hospital death | Outcome |

The first column represents the numerical identifier for each state, following the state name identifier.

We performed an exhaustive search within the group of AECOPD patients extracted from MIMIC-III for antibiotics used to treat COPD. We then extracted information regarding administration times of those antibiotics for the subset of patients by preparing a comprehensive list of antibiotics names and codes from the National Drug Codes (NDC) directory^24^ and the National Library of Medicines RXNorm database.^25^ The antibiotics used to treat these patients were: *doxycycline, azithromycin, levofloxacin, oseltamivir, vancomycin, trimethoprim, fluoroquinolones, vibramycin, ofloxacin, clarithromycin, telithromycin, amoxicillin, cefuroxime, pipericillin-tazobactam, cefepime*. Data extraction and preprocessing was done via Structured Query Language (SQL) queries and Python scripts. This process included the removal of duplicate and implausible data entries. For example, if a time-stamp for a clinical measurement occurred after a patient was discharged, that patient record was excluded.

### Markov Chain Model

The Markov chain model consisted of three mutually exclusive (transient) health states: no acute respiratory failure (NARF), acute respiratory failure type 1 (ARF_1_), and acute respiratory failure type 2 (ARF_2_), and two (absorbing) outcome states: discharge and death (Figure 2). The patient histories, *H* = {*h*_0_, *h*_1_, …, *h*_*n*_}, were represented by a set of state transition events, {(*s*_0_, *t*_0_), …, (*s*_*n*_, *t*_*n*_)}. Here a time-stamp, *t* denoted the exact time a patient entered a particular state, *s*. We used this information to compute transitions as a function of time. For every sequential pair of transition events, we calculated the actual time between them in hours as (*s*_*i*+1_ − *s*_*i*_). We aggregated all patient histories to construct the state transition matrix *T*, where *x*_*ij*_ denotes the total number of hours spent in state *i* before moving to state *j*.

**Figure 2:**
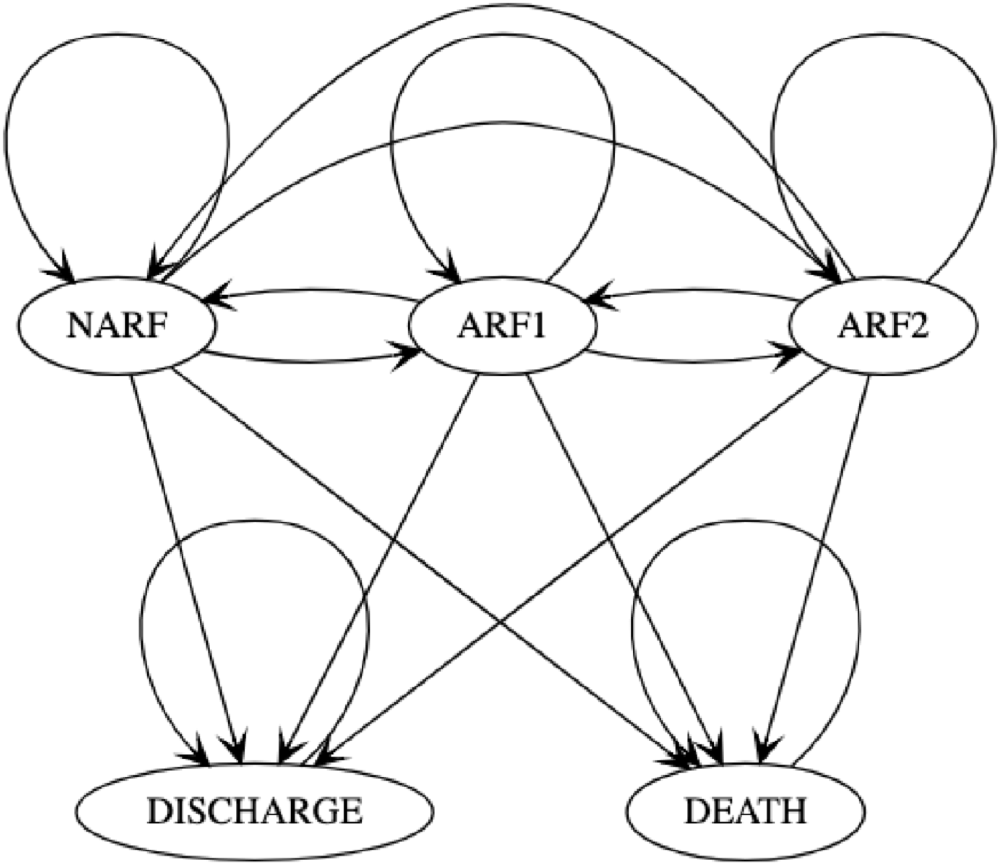
A 5-state first-order Markov model for AECOPD.

Transition probabilities matrices (TPMs) for the Markov model were estimated using the ratio of the number of hours spent in a specific state before transitioning over the total number of hours spent in that state. Each transition probability (or model parameter) represents the estimated likelihood of a patient changing from one state to another during their hospital stay. Specifically, we calculated transition probabilities by:

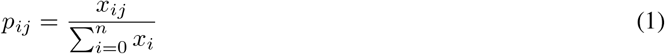

where *p*_*ij*_ is the probability of transitioning from state *i* to state *j* at any given time. We set *p*_*ij*_ equal to the percentage of hours spent by individuals in state *i* and ending in state *j* relative to the total amount of time spent in state *i*. Note that *p*_*ii*_ = 1 for an outcome state. The model has a corresponding initial probability vector that is calculated from the initial percentage of patients starting in each of the health states.

### Monte Carlo Simulation

Monte Carlo simulations were performed to estimate the outcomes from antibiotic administration on AECOPD patients in the ICU. We simulated the transitions of individual patients over time until they were either discharged or they died for the following two questions of interest:

i. What is the impact of antibiotics on death for hospitalized AECOPD patients?
ii. What is the initial timing of antibiotics for hospitalized AECOPD patients and the impact on death?

We ran the simulation to compare the in-hospital death rate for two populations, those that received antibiotics and those that did not. Patients were classified into four groups, ALL (the full population), ANTIBIOTICS (those for whom antibiotics were administered), and NO ANTIBIOTICS (those who did not receive antibiotics). For each of the two questions, the simulation was run for 100 replications (or cohorts), each of size 1000 patients. Each patient was probabilistically assigned to an initial health state based on the initial state probabilities. The occurrence of transitions between transitioning between two states was based on the transition probabilities from the possible range of values, specified in the *P* matrix. Because the probability of a patient leaving an outcome state is zero, once a patient entered an outcome state, the process ended and a new patient was simulated. The mean, minimum, and maximum of the patient outcomes for the 100 replications was calculated and used for simulation statistics and model validation. In order to validate the simulation, we obtained a baseline from the raw EHR data and compared the observed outcomes (mean and variance) to estimated results from simulation.

Sensitivity analysis was conducted to quantify the effects of changes in model parameters (i.e., transition probabilities). A small change randomly chosen between plus or minus 10% of each transition probability value was applied to the model parameters. That is:

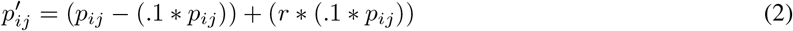

and normalized using the row sum for each state.

A new TPM was obtained by normalizing the generated values so that the row sums equalled 1. The outcomes were then re-computed from the simulation using the new TPM.

## RESULTS

Table 2 describes the characteristics of the patient groups, including the percentage of patients who received antibiotics. Notably, the NO ANTIBIOTICS group had the highest death rate (9.88%) and 0% of the patients were administered antibiotics. Figure 3 shows the transition probabilities and initial state probability vectors for each group, which derived from the Markov chain models created from the raw data. Additionally, figure 3 includes an additional TPM for a group, HALF, which was computed using the TPMs from the ANTIBIOTICS and NO ANTIBIOTICS group. The HALF TPM was used to conduct an analysis for administering 50% of the population antibiotics. We used the initial state probabilities from the ALL group to simulate each of the four models.

**Table 2:** Descriptive characteristics of patients included.

|  | ALL<br>(*n* = 697) | ANTIBIOTICS<br>(*n* = 181) | NO ANTIBIOTICS<br>(*n* = 516) |
| --- | --- | --- | --- |
| Age in years <sup>a</sup> | 70.81 ± 11.18 | 70.6 ± 10.76 | 70.88 ± 11.34 |
| Women <sup>b</sup> | 357 (51.22%) | 91 (50.28%) | 266 (51.55%) |
| Death <sup>b</sup> | 59 (8.46%%) | 8 (4.41%%) | 51 (9.88%%) |
| LOS, days <sup>c</sup> | 8.63 | 6.92 | 9.26 |
| Administered antibiotics | 25.9% | 100% | 0% |
| Initial antibiotics |  |  |  |
| Drug name |  |  | - |
| *vancomycin* | 58 (8.32%) | 58 (32.04%) | - |
| *azithromycin* | 47 (6.59%) | 47 (25.97%) | - |
| *levofloxacin* | 38 (5.45%) | 38 (20.99%) | - |
| Administration, hours <sup>c</sup> | 27.37 | 27.37 | - |

<sup>a</sup> mean ± standard deviation<sup>b</sup> count (%)<sup>c</sup> mean **Figure 3:** Model parameter estimations for all AECOPD patients administered antibiotics and not administered antibiotics are presented by group. Row and column labels correspond to health and outcome states in Table 1.

**Figure 3:**
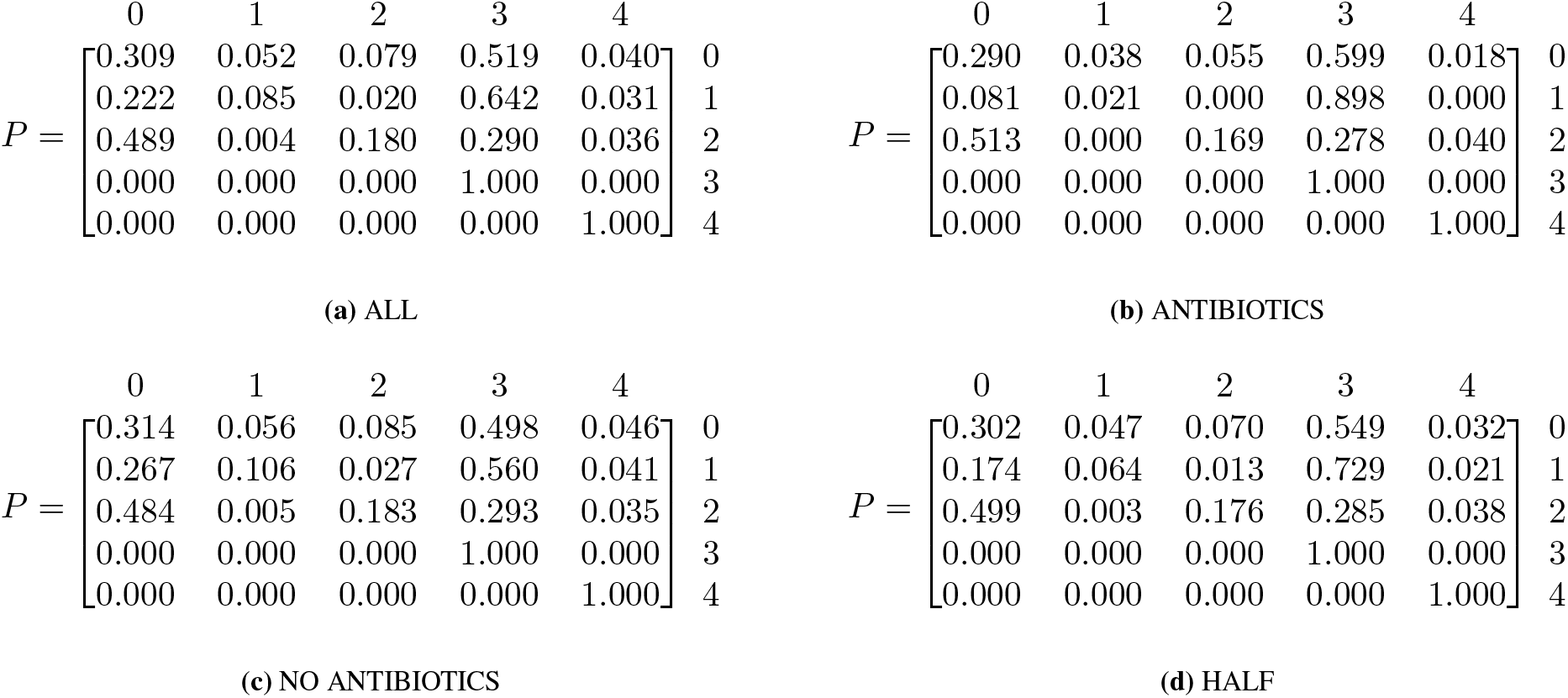
Model parameter estimations for all AECOPD patients administered antibiotics and not administered antibiotics are presented by group. Row and column labels correspond to health and outcome states in Table 1.

Figure 4a shows the estimated percentage of ICU patients with AECOPD that died as a function of the percentage that received antibiotics. Note that the actual number of patients that received antibiotics in the EHR was 25.9% with a death rate of 8.46% for the full cohort. This falls within the death rate interval [5.3%, 9.6%] estimated by the simulation. Also note that as the number of patients administered antibiotics increased, the percentage of deaths significantly decreased. Specifically, when all patients were given antibiotics, the death rate decreased by 50% vs not administering antibiotics.

**Figure 4:**
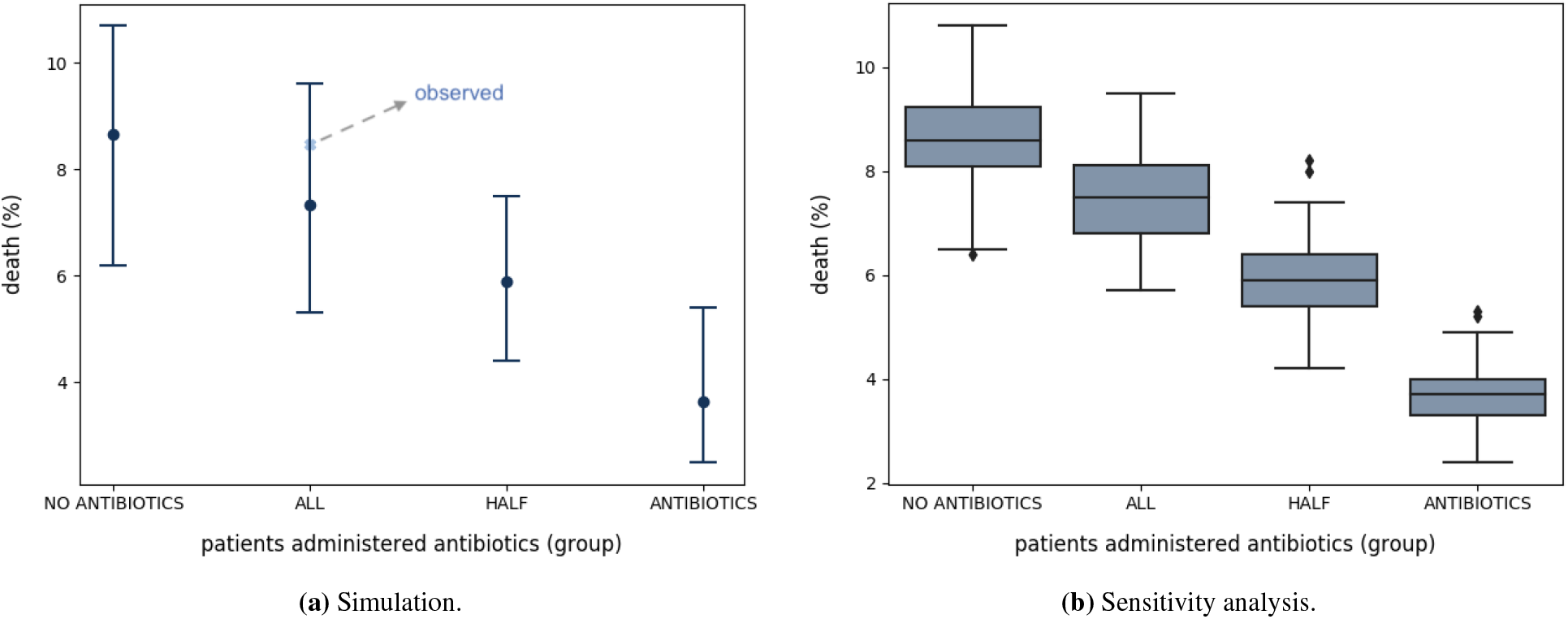
Results for **(a)** The estimated percentage of death for AECOPD based on antibiotics or no antibiotics administration. The x-axis is ordered by the percentage patients administered antibiotics and labeled by group. The y-axis represents the percentage deaths for that particular percentage of patients. The top and bottom horizontal lines are the maximum and minimum percentage of patient deaths, respectively. The black data point represents the average percentage of patient deaths, and the data point labeled, *observed*, represents the actual percentage of AECOPD patients that died when administered antibiotics. **(b)** The death percentage estimated for sensitivity analysis. Outliers are represented by data points. The median death percentage is represented by the horizontal line inside of the box. Top and bottom horizontal lines represent values higher than and the median.

The average time to antibiotic administration was 27 hours, and 32% of ANTIBIOTICS patients were administered *vancomycin* as the initial antibiotic (Table 2). Models were created using data from patients that were given antibiotics within the following specified time-frames: i) within 6 hours of hospital admission, ii) between 6 and 24 hours of admission, iii) between 24 and 48 hours of admission, and iv) after 48 hours of admission. We also used the initial state probabilities from the ALL group to simulate each of the four models.

Interestingly, Figure 6a shows a 5.5% mortality in the group of cohorts that received antibiotics after 48 hours vs 1.8% in the group that received antibiotics between 24 and 48 hours. The sensitivity analyses was performed by the perturbation of each probability in the transition probability matrix, and then observing how that perturbation changed the outcome (death). Figure 6b shows the results of the sensitivity analysis for different groups.

**Figure 5:**
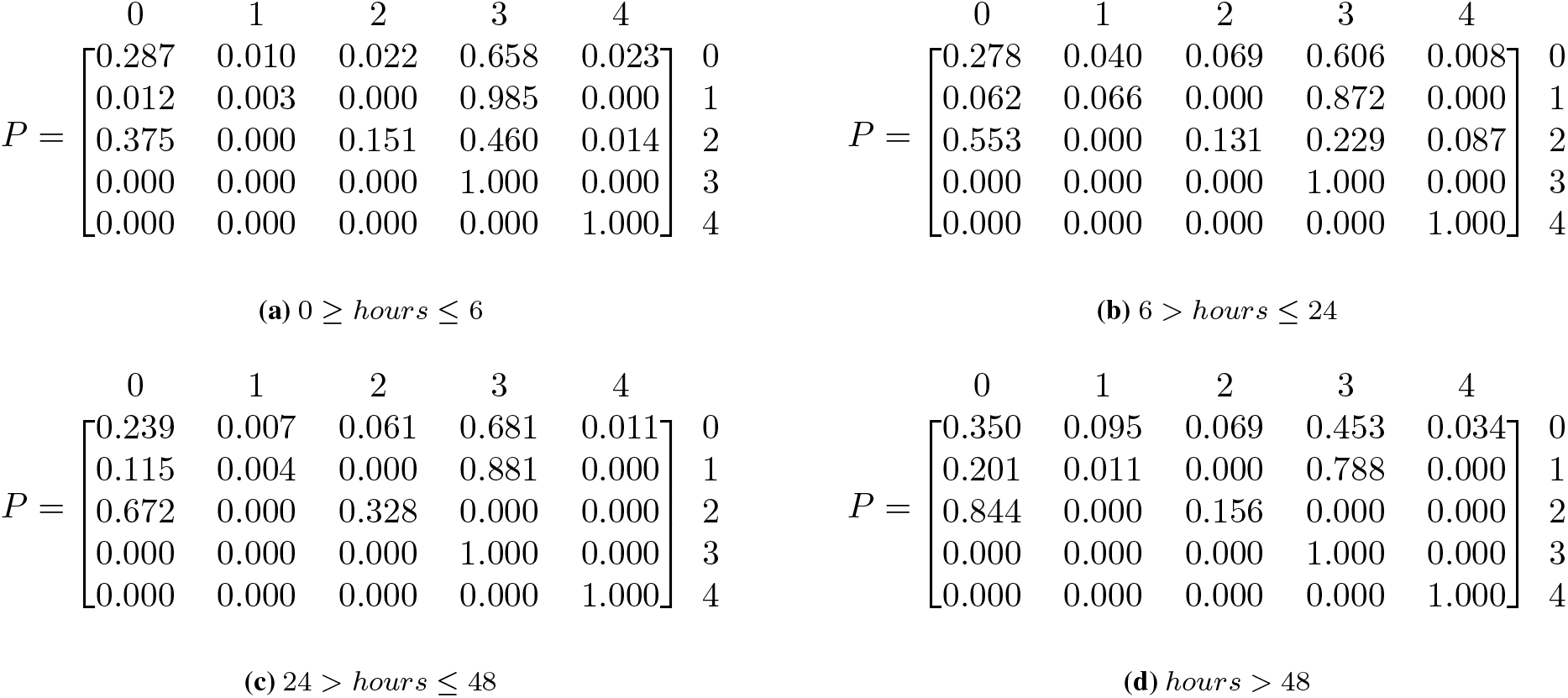
Model parameter estimations for AECOPD patients given antibiotics according to initial timing administered in hours are presented by group. Row and column labels correspond to health and outcome states in Table 1.

**Figure 6:**
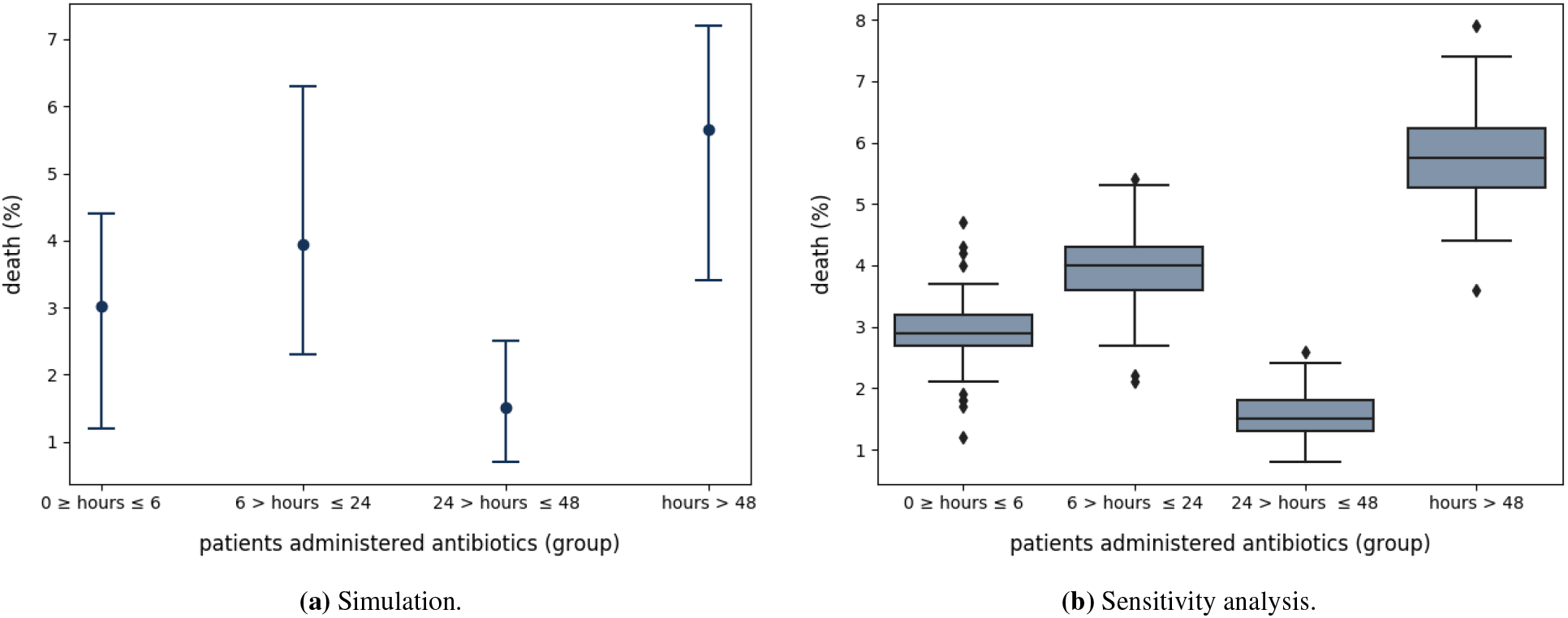
Results for **(a)** The estimated percentage of death for ICU patients with AECOPD based on the initial timing antibiotics were administered. The x-axis represents the time from admission in which patients were administered antibiotics, labeled by group. The y-axis represents the percentage deaths for that particular percentage of patients. The top and bottom horizontal lines are the maximum and minimum percentage of patient deaths, respectively. The black data point represents the average percentage of patient deaths, and the data point labeled, *observed*, represents the actual percentage of AECOPD patients that died when administered antibiotics. **(b)** Outliers are represented by data points. The median death percentage is represented by the horizontal line inside of the box. Top and bottom horizontal lines represent values higher than and the median.

## DISCUSSION

Physicians frequently make critical decisions in the face of uncertainty,^3^ weighing the risks and benefits of patient treatments and outcomes. Uncertainty is inevitable in medical reasoning and decision making, especially in critical care settings where the complete patient history may not be available. We introduced an approach to systematically capture, analyze, and estimate the effect of treatment interventions on outcomes through modeling and estimating uncertainties using EHR data. A major contribution of our work is the ability to derive transition probabilities from empirical time-series data by scaling the probabilities over time. This approach captures the duration of time that a patient spent in a particular state and allows extraction of temporal inferences from mathematical models using realworld data. Additionally, we demonstrated our approach to, for the first time, asses the administration and timing of antibiotics for AECOPD and the corresponding impact on in-hospital mortality. Our findings support a general approach of estimating patient outcomes from raw clinical data.

Chronically ill patients are susceptible to frequent changes in health status,^26^ especially when under care in an ICU. For AECOPD patients in the ICU, clinical indicators can change suddenly, causing a patient to move between the health states repeatedly, as illustrated in Figure 2. In our study we have seen an overall reduced mortality rate (Figure4a) in patients with AECOPD admitted to the ICU who received antibiotics in comparison to those who did not. These findings are consistent with previous evidence that supports the use of antibiotics in critically ill patients given they frequently have community acquired pathogens.^27^ Antibiotics were shown to reduce all-cause mortality in critically ill patients and also reduce treatment failure after 4 weeks of discharge.^28^ Our findings suggest that there may be a reduction in mortality rate when initiation of antibiotics occurs earlier in settings of AECOPD patients with severe respiratory failure warranting an ICU admission.

### Limitations

One limitation of our study is that all of our data was limited to that in the ICU. Additional patient history might have helped to refine the findings. Another limitation is the process of selecting patient cohorts. The retrospective EHR data used required modeling patients who were assigned an AECOPD ICD-9 code. Though ICD-9 codes have been used for identifying patients when conducting retrospective clinical studies, accurately identifying COPD patients can be difficult in the absence of patient medical history.^23^ COPD is diagnosed by lung function tests and staged according to the progression of the disease. Thus, simply using ICD-9 codes may not be enough information to identify an individual with COPD but used identify an individuals with COPD-like symptoms (e.g., wheezing, or noisy breathing, chest pain) who was given a COPD ICD-9 diagnosis. Such symptoms may be documented as free-text, unstructured data in EHR clinical notes. We also acknowledge limitations with modeling EHR data as Markov chains in that state transitions in first-order Markov chains only depend on the current state. Thus, such modeling does not account for confounding factors that previously impacted a patient prior to entering their current state. While Markov models are valuable when history is not important or available, they potentially lack accuracy when history matters.^29^ Work has been done^29^ to develop more complex Markov chain modeling methods that incorporate history.

### Future Work

The development of clinical data networks such as Patient-Centered Clinical Research Network (PCORnet)^30^ and Informatics for Integrating Biology and the Bedside (i2b2),^13^ allow for the integration of various clinical and research data sources into a single repository, increasing the amount and availability of clinical data. These platforms offer tools for cohort discovery and analytics that are currently being used for clinical research. Specifically, in addition to using ICD-9 codes, i2b2’s natural language processing (NLP) feature for cohort discovery can be used for identifying AECOPD patient by extracting medications, smoking status, and diagnosis from clinical notes. While these tools allow simple data analysis, they do not offer support for granular temporal data analysis. Our methods can be integrated into such existing systems to allow more complete analysis.

Our methodology can also supplement real-time analysis of physiological data streams^31^ in hospital settings and be applied to both common and more complex clinical conditions. That is, we defined health states for our model that encompassed several clinical variables. However, given the vast space of clinical conditions and corresponding clinical criteria that defines the conditions, some health states may not be well represented. For example, in our state space NARF is defined as no acute respiratory failure, which means that the only criteria that determines if a patient is in NARF is that the constraints used to define ARF_1_ and ARF_2_ are not met. In this case, NARF does not reveal much information about the condition of the patient. To address this, information from various data sources such as past research studies,^8^ clinical trials,^32^ and medical devices can be combined to define additional clinical states. Clinical data warehouses^12,13,30^ have proven to be valuable for overcoming the barriers of data access and availability as they provide access and structure of medical data from multiple sources as well as tools for clinical discoveries through data analysis. Our work can be integrated as an API for platforms such as i2b2^13^ for complex data analytics and generalized to fit clinical data standards such as Fast Healthcare Interoperability Resources (FHIR)^33^ for integration with healthcare systems.

## CONCLUSION

Early administration of antibiotics for AECOPD patients in ICU settings can potentially reduce mortality rate. Such evidence can be generated for clinical guidelines and more generally, clinical discoveries, by developing comprehensive probabilistic modeling and simulation techniques that integrate temporal aspects of EHR data. As clinical researchers continue to strive to achieve data standardization, interoperability, and reproducibility, the methods presented in this paper can help leverage multiple EHR data sources and technological advancements.

## Data Availability

"MIMIC-III (Medical Information Mart for Intensive Care III) is a large, freely-available database comprising deidentified health-related data associated with over forty thousand patients who stayed in critical care units of the Beth Israel Deaconess Medical Center between 2001 and 2012.”

https://mimic.physionet.org/

